# Preventable road deaths in 72 countries, 2021

**DOI:** 10.64898/2026.01.29.26345165

**Authors:** Leon S. Robertson

## Abstract

World Health Organization recommendations to reduce road deaths were examined to assess the potential reductions that could be realized in countries that have not adopted them. Data from 72 countries on recommended speeding laws, alcohol laws, and vehicle safety standards were analyzed, controlling statistically for differences in average temperatures and population density per square kilometer. Using regression coefficients, estimates of the reductions that would be realized if each countermeasure were adopted in countries not currently employing it were calculated. The coefficient on alcohol laws was not significant, but deaths in these countries would likely decline by about 23 percent if speeding laws were improved. The road death would have been about 55 percent lower if vehicle safety standards for imported vehicles had been adopted. New and used vehicles that did not adhere to the standards were sold in low-income countries. Better data identifying clusters of specific collision types (pedestrians in the dark, animals, fixed objects) could lead to the adoption of countermeasures known to be effective.

## Introduction

The effectiveness of various measures to reduce road deaths has been known for decades. Researchers have repeatedly found that standards for improved vehicle energy absorption in crashes and crash avoidance technology, such as electronic stability control, laws requiring seat belt use by closed-vehicle occupants and helmet use by riders of open vehicles, and legal limits on speeding and drivers’ blood alcohol concentration, are associated with reduced deaths [1-6]. In 2004, the World Health Organization (WHO) included these measures as recommendations for all countries [7]. By 2020, the WHO reported that most countries required seatbelt use for front-seat occupants and helmets for open-vehicle riders, but the other recommendations continued to be neglected in many countries. Globally, the road death rate per 100,000 vehicles declined from approximately 18 to about 15 between 2010 and 2020, but increased in 2021 [8]. The report noted that in 2021, lower-income countries had substantially higher road death rates per capita, but not inevitably. In Africa, lower-middle-income countries had higher death rates than the lowest-income countries.

Failure to adopt standards for vehicle crash avoidance and crashworthiness allows manufacturers to sell vehicles with inferior protection. Major manufacturers sell new vehicles in low- and middle-income countries that do not meet safety requirements, and some models differ from the same models sold in their home countries [9-10]. Furthermore, high-income countries reduced the number of older vehicles that did not meet safety and pollution standards by shipping them to low-income countries. [11] Safer vehicles, helmets, and fines for speeding and alcohol violations impose higher costs on road users, but the costs of injuries and deaths exceed those costs. [12] Researchers at the World Bank and elsewhere indicate that reductions in road injuries enhance economic growth in low- and middle-income countries. [13-14]

Road deaths are also associated with other factors such as ambient temperature and the number of people per square kilometer (density). Road deaths are associated with warmer weather, as some people avoid traveling in colder weather. [15-16] Lower road death rates are also associated with increased population density per square kilometer. [17]

The purpose of the study reported here is to estimate the death reductions that would have occurred if countries that neglected the WHO recommendations had complied with them, adjusted for temperature and population density. The results are limited to the 72 countries for which all data were available, but the percentage reductions should apply to others.

## Materials and Methods

The research plan was to match road deaths in 2021, the latest year available, to WHO assessments of the status of the mentioned countermeasures and indicators of other risk factors for as many countries as data availability allowed. Only four countries did not have a seatbelt use law, and six did not have a helmet use law, so those countermeasures were not included.

Most road death research uses Poisson or Negative Binomial regression to analyze data, but these models require population or vehicles as an offset variable. In this study, the results of such models would be unduly influenced by China and India, whose combined populations are 52 percent of the populations of the 72 countries studied. Therefore, a least squares regression model was used to estimate the contribution of each countermeasure and risk factor. The form of the equation is:

Log (deaths/100,000 population) = a + b_1_(alcohol BAC law) + b_2_(speeding law) + b_3_(vehicle safety regulations) + b4(log(average temperature))+ b_5_(log((population density)) + e

The logarithm of the death rates, temperature, and density was used to reduce skewness in the distributions. The recent claim that road deaths per capita are higher in low and high-income countries than in middle-income countries [18] is not true of the countries in this study. An index of economic purchasing power was considered for inclusion, but after viewing the correlation coefficients presented in the results, it was dropped from the model because it was too highly correlated with other variables and was substantially controlled indirectly by the inclusion of those variables.

To estimate the number of deaths that would have occurred if countries that did not apply countermeasures had done so, the model coefficients were applied to each country, and the expected deaths that would have occurred with full implementation of countermeasures were summed. Data were available for 72 countries with a total population of 5.4 billion people and 779,161 road deaths in 2021.

The data sources and their references are listed in Table 1. The alcohol and speeding laws were coded 1 if they met WHO’s criteria and 0 otherwise. The vehicle safety standards were the number adopted, ranging from 0 to 8. To account for variation in the temperature experienced by people in different regions of a country, the average temperature was calculated by summing the population in each region of the country times the temperature in that region and dividing the total by the country’s total population.

**Table 1.**
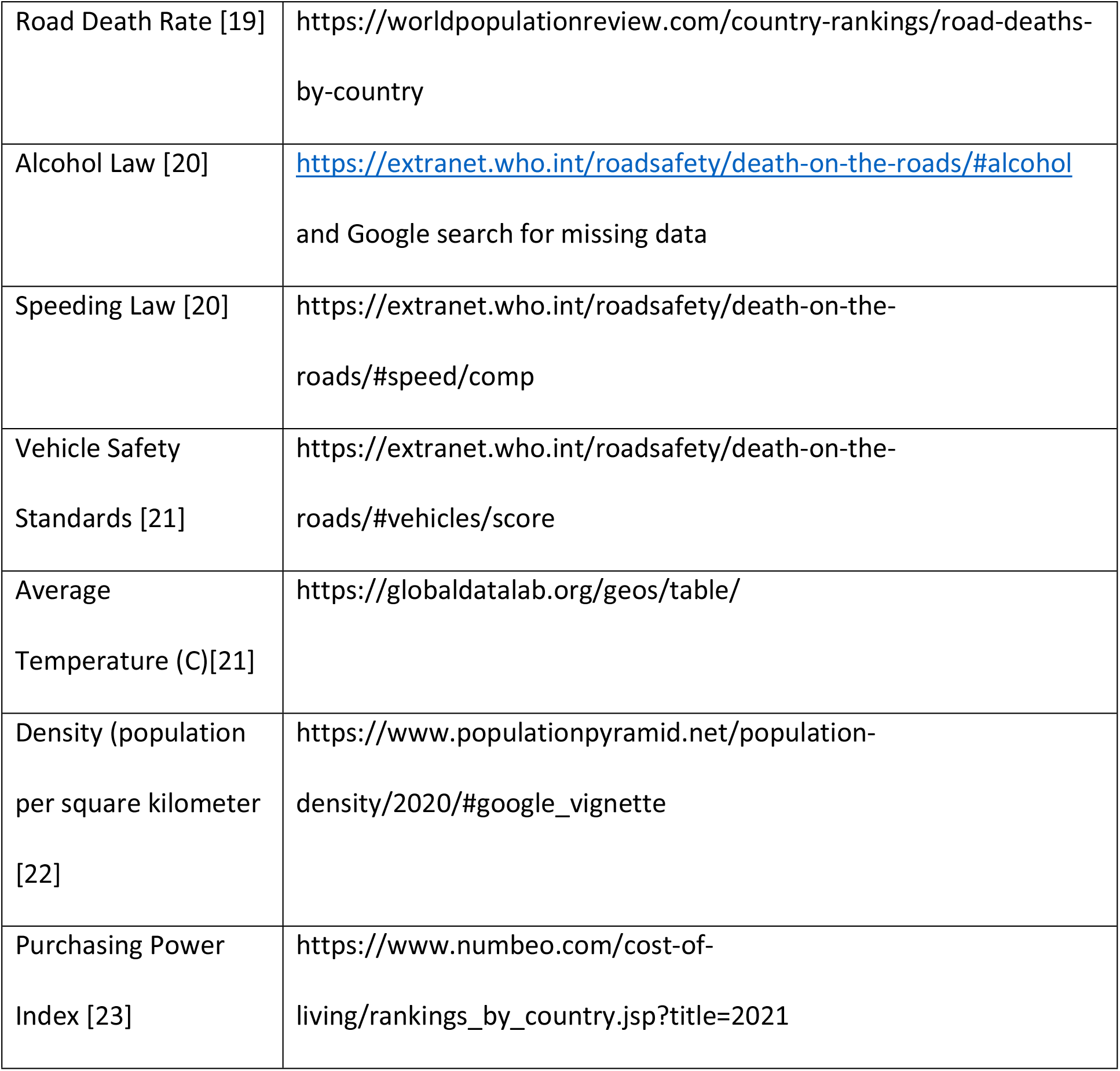
References to data sources.

|  |  |
| --- | --- |
| Road Death Rate [19] | https://worldpopulationreview.com/country-rankings/road-deaths-by-country |
| Alcohol Law [20] | https://extranet.who.int/roadsafety/death-on-the-roads/#alcohol<br>and Google search for missing data |
| Speeding Law [20] | https://extranet.who.int/roadsafety/death-on-the-roads/#speed/comp |
| Vehicle Safety Standards [21] | https://extranet.who.int/roadsafety/death-on-the-roads/#vehicles/score |
| Average Temperature (C)[21] | https://globaldatalab.org/geos/table/ |
| Density (population per square kilometer [22]) | https://www.populationpyramid.net/population-density/2020/#google_vignette |
| Purchasing Power Index [23] | https://www.numbeo.com/cost-of-living/rankings_by_country.jsp?title=2021 |

## Results

The means and standard deviations of the included variables are shown in Table 2. The large standard deviations relative to the means indicate the diversity of the countries included. Robust standard errors were employed in the regression analysis to reduce heteroskedasticity.

**Table 2.**
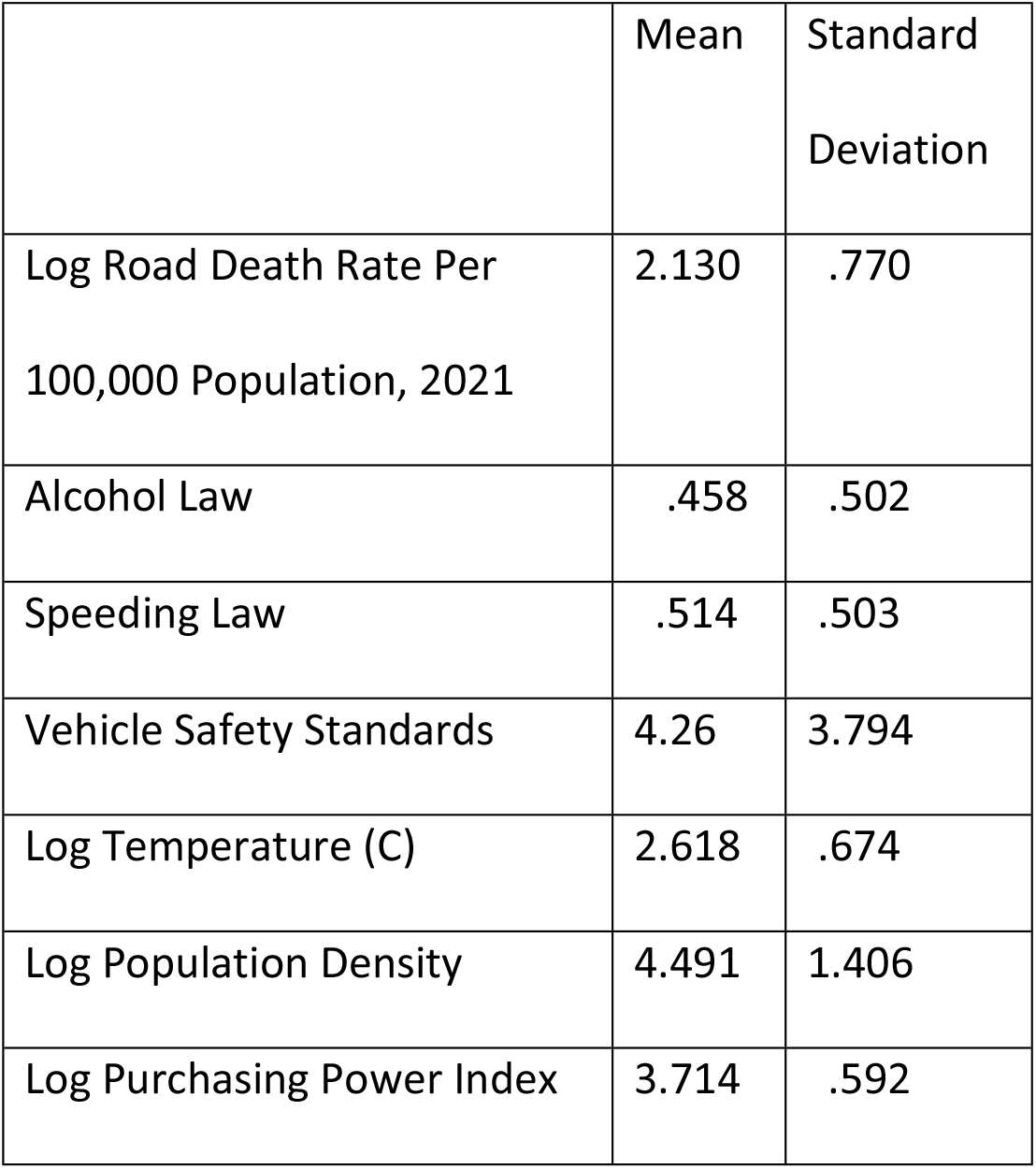
Means and Standard Deviations of Included Variables.

|  | Mean | Standard<br>Deviation |
| --- | --- | --- |
| Log Road Death Rate Per<br>100,000 Population, 2021 | 2.130 | .770 |
| Alcohol Law | .458 | .502 |
| Speeding Law | .514 | .503 |
| Vehicle Safety Standards | 4.26 | 3.794 |
| Log Temperature (C) | 2.618 | .674 |
| Log Population Density | 4.491 | 1.406 |
| Log Purchasing Power Index | 3.714 | .592 |

Table 3 presents the least squares correlations among the noted variables. Without correction for other risk factors, the road death rate per 100,000 population is lower in countries with better alcohol and speeding laws, more vehicle safety standards, and higher purchasing power. It is higher in countries with higher average temperatures, but not correlated with population density. Because the correlation of purchasing power with each of the predictor variables is relatively high, it was not included in the regression analysis to avoid distorting the coefficients.

**Table 3.** Least squares correlations of the country road death rates with potential predictor variables, and among them, 73 countries, 2021.

|  | Alcohol<br>Law | Speed<br>Law | Vehicle<br>Standards | Log<br>Temperature | Log<br>Density | Log<br>Purchasing<br>Power |
| --- | --- | --- | --- | --- | --- | --- |
| Log Death<br>Rate | -.36 | -.42 | -.66 | .52 | -.05 | -.74 |
| Alcohol Law |  | .28 | .45 | -.25 | -.08 | .48 |
| Speed Law |  |  | .37 | -.13 | .03 | .41 |
| Vehicle<br>Safety<br>Standards |  |  |  | -.43 | .07 | .77 |
| Log Average<br>Temperature |  |  |  |  | .53 | -.49 |
| Log<br>Population<br>Density |  |  |  |  |  | -.02 |

Usually, data of this type are analyzed using age-standardized rates. Age-standardization assumes that the risk to each age group is similar among the units, in this case, nations, being studied. That is unlikely for road deaths. In an attempt to age-standardize the death rate, huge differences in the age distributions among countries were evident. The countries with a larger proportion of children less than 10 years old tended to have substantially higher death rates (Figure 1). In the US, that is the age group with the lowest rate [24]. If that is true in countries where children are the largest proportion of the population, the death rates in older age groups are huge by comparison to other countries. An attempt to use the US rates by age in 2021, to standardize the rates of countries in the study, produced a model substantially inferior in goodness of fit to ordinary least-squares regression of the logged unadjusted rates.

**Figure 1.**
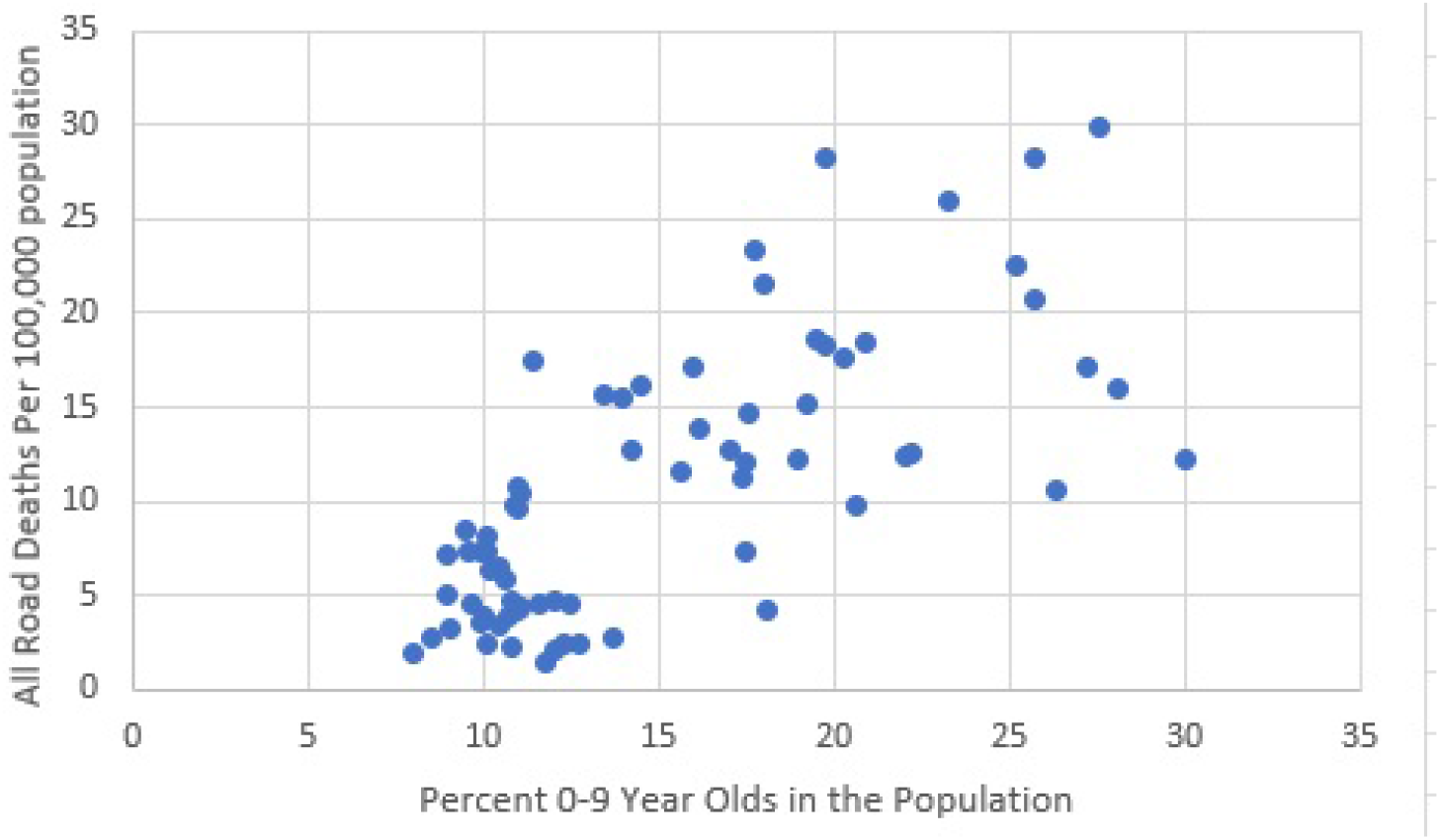
Death Rates Per 100,000 Population by Percent of 0–9-Year-olds in the Population.

The regression coefficients, robust standard errors, and odds ratios are in Table 4. Lower death rates are associated with better speeding and alcohol laws, but the possibility of random variation cannot be ruled out at the conventional 95 percent confidence intervals in the case of alcohol laws. More vehicle safety standards predict lower death rates. Higher temperatures are associated with a higher risk, and higher population density with a lower risk.

**Table 4.** Log Road Deaths Per 100,000 Population: regression coefficients, standard errors (SE), and odds ratios, 2021.

|  | Regression<br>Coefficients<br>(Robust<br>Standard Errors) | Odds Ratio | OR 95%<br>Confidence<br>Intervals |
| --- | --- | --- | --- |
| Speed Law | -.267 (.118) | .766 | .605, .969 |
| Alcohol Law | -.049 (.125) | .952 | .743, 1.220 |
| Vehicle<br>Standards | -.095 (.020) | .909 | .874, .946 |
| Log<br>Temperature | .483 (.113) | 1.621 | 1.294, 2.031 |
| Log Density | -.141 (.049) | .868 | .787, .958 |
| Intercept | 2.060 |  |  |
| R-squared | .64 |  |  |

The predicted deaths when countermeasures were set to the best WHO ratings in countries that had not fully adopted them are shown in Table 5. The sum of the predicted deaths across the countries indicated a 3.9 percent overprediction in the aggregate, a reasonably good fit of the model. Applying the coefficients to each country and setting the speed law to 1, and separately setting the vehicle standards to 8, indicated that improving speed laws in countries where the WHO indicates them as less than optimal would have reduced deaths by 23.4 percent. If the 44 countries that had fewer than the eight safety standards had those standards, the estimated number of deaths would have been about 54.8 percent lower. If the average temperature in these countries increases by one degree Celsius, a 2.7 percent increase in deaths is predicted.

**Table 5.**
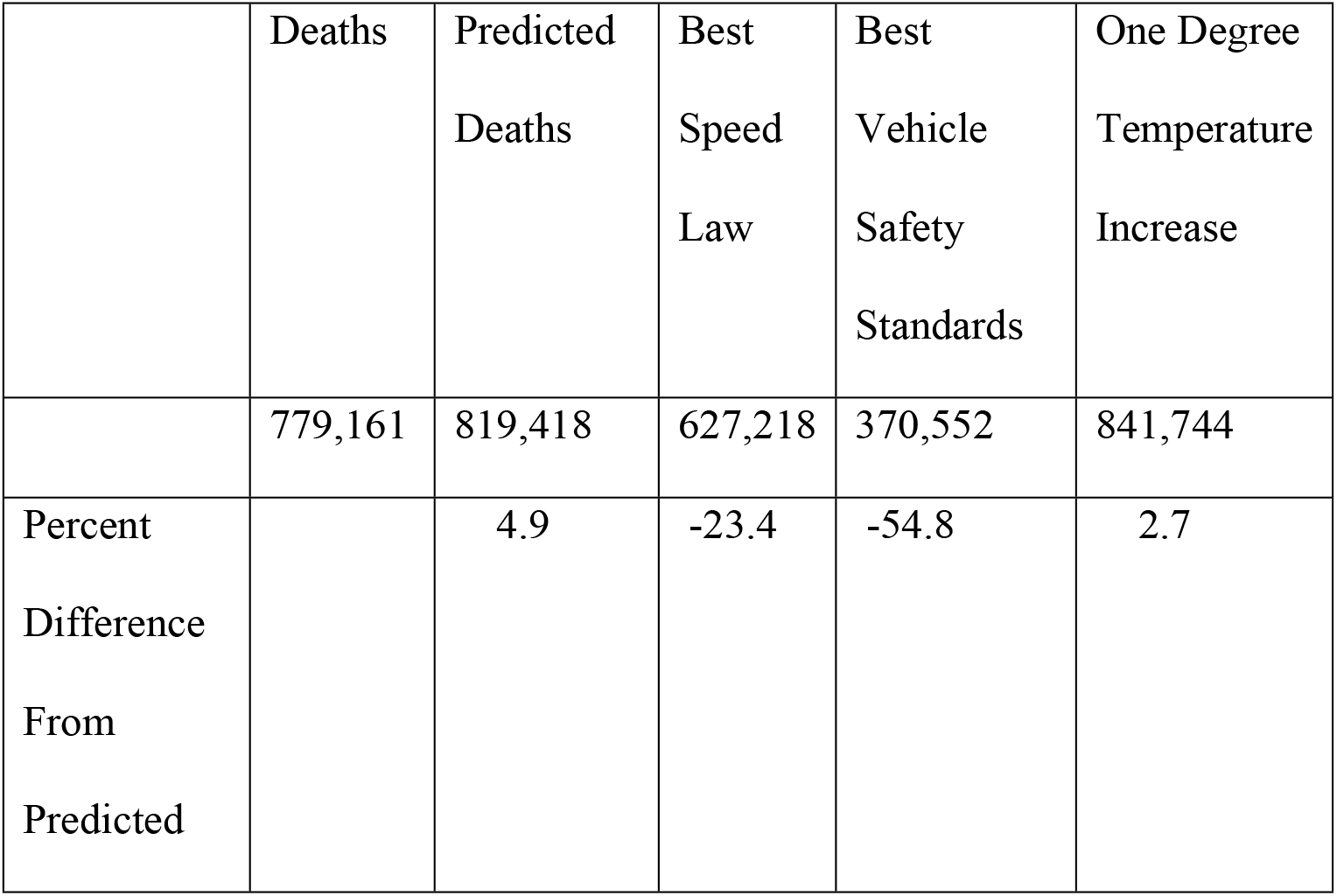
Predicted deaths if speeding laws and vehicle standards were the best.

|  | Deaths | Predicted Deaths | Best Speed Law | Best Vehicle Safety Standards | One Degree Temperature Increase |
| --- | --- | --- | --- | --- | --- |
|  | 779,161 | 819,418 | 627,218 | 370,552 | 841,744 |
| Percent Difference From Predicted |  | 4.9 | -23.4 | -54.8 | 2.7 |

## Discussion

The data indicate that adopting vehicle safety standards and improving speeding laws are likely to lower death rates. Alcohol laws are more problematic. A police officer cannot judge a driver’s blood alcohol level without stopping them and doing a test. The alcohol laws do not cover inebriated pedestrians who are at substantial risk. One of the earliest case-control studies of alcohol’s role in road safety compared the blood alcohol of fatally injured pedestrians to that of people stopped while walking at the same time of day and day of week. The fatally injured were three times as likely to have a blood alcohol level of.05 percent or more by weight as the controls. [25] Publicized efforts to enforce alcohol laws sometimes result in mortality reductions, but the results are temporary. [26-27] Speed can be measured remotely, and speeding laws are more likely to be enforced than alcohol laws, but drivers sometimes signal other drivers of enforcement ahead.

Allowing cheaper new and used vehicles that are not subject to safety standards in a country may be politically popular, but when the vehicles do not conform to safety standards, the price in premature loss of life is severe. The lack of standards is concentrated in countries with low purchasing power. Countries with moderate purchasing power that require all vehicle safety standards include Bulgaria, Croatia, Cyprus, Estonia, Greece, Hungary, Italy, Latvia, Lithuania, Poland, Portugal, Romania, Slovenia, and Spain – all European Union countries with common standards.

If countries without vehicle safety standards adopt them, the replacement of less-safe vehicles will occur over an extended time. Countermeasures more effective immediately are available. Targeted interventions based on data showing clusters of specific types of road deaths and severe injuries have been found effective, such as lighting sections of road where pedestrians were struck at night, removing livestock from roads, and placing energy-absorbing barriers or barriers sloped to guide vehicles back onto roads where vehicles frequently leave roads, among many others. [28-30] To maximize the effectiveness of such measures, data on where, how, and when fatalities are occurring is needed. [31] Local public health workers, police, or emergency responders can collect such data and use pin maps with different colored pins to indicate pedestrian vs. occupant and daylight vs. darkness. Choice of a countermeasure is often evident from a visit to sites where clusters are found. Charitable organizations are more likely to fund projects with data-supported prospects for success. [13]

## Data Availability

The data are available at: https://dataverse.harvard.edu/dataset.xhtml?persistentId=doi%3A10.7910%2FDVN%2FUZYMIF&version=DRAFT

## References

1. Foldvary LA, Lane JC. The effectiveness of compulsory wearing of seat-belts in casualty reduction (with an appendix on chi-square partitioning-tests of complex contingency tables). Accident Analysis & Prevention. 1974. 6(1):59–81.

2. Robertson LS. Reducing death on the road: the effects of minimum safety standards, publicized crash tests, seat belts, and alcohol. American Journal of Public Health. 1996.86(1):31–34.

3. Peng Y, Vaidya N, Finnie R, Reynolds J, Dumitru C, Njie G, et al. Universal Motorcycle Helmet Laws to Reduce Injuries: A Community Guide Systematic Review. American Journal of Preventive Medicine. 2017 Jun;52(6):820–832.

4. Karkhaneh, M, Kalenga KC, Hagel BE, Rowe BH. Effectiveness of bicycle helmet legislation to increase helmet use: a systematic review. Injury Prevention. 12:76–82, 2006. 10.1136/ip.2005.010942.

5. Mann RE, Macdonald S, Stoduto G, Bondy S, Jonah B, Shaikh A. The effects of introducing or lowering legal per se blood alcohol limits for driving: an international review. Accident Analysis & Prevention. 2001 Sep;33(5):569–83.

6. Andreuccetti G, Carvalho HB, Cherpitel CJ, Ye Y, Ponce JC, Kahn T, et al. Reducing the legal blood alcohol concentration limit for driving in developing countries: a time for change? Results and implications derived from a time-series analysis (2001-10) conducted in Brazil. Addiction. 2011 Aug 23;106(12):2124–31.

7. Peden M, Scurfield R, Sleet D et al. (Eds.) World Report on Road Traffic Injury Prevention. Geneva, Switzerland: World Health Organization, 2004.

8. Global status report on road safety 2023. Geneva, Switzerland: World Health Organization, 2023. https://iris.who.int/server/api/core/bitstreams/46275f9f-ef66-4892-8ddd-a496ef8c1b74/content

9. Ward D, Truong J. Global enhancement of vehicle safety - the urgency of now. Journal of the Australasian College of Road Safety. 2016 27:48–51.

10. Campbell T. 7 Popular auto companies still using outdated safety standards overseas. Newsbreak, May 25, 2025. https://www.newsbreak.com/clever-dude-288519147/4023301654918-7-popular-auto-companies-still-using-outdated-safety-standards-overseas

11. United Nations Environmental Programme. Used Vehicles and the Environment: A Global Overview of Used Light Duty Vehicles, Flow, Scale and Regulation. 2020. https://wedocs.unep.org/items/c3860caa-7c37-4c6f-92c9-5c269d883760/full

12. Byaruhanga CB, Evdorides H. The impact of indirect benefits (reduced travel time, fuel use and emissions) in cost benefit analysis of road safety countermeasures. Traffic Injury Prevention. 2024 Mar 5;25(3):434–9. 10.1080/15389588.2024.2322665

13. World Bank Group. The High Toll of Traffic Injuries: Unacceptable and Preventable. Washington DC, 2017. https://openknowledge.worldbank.org/server/api/core/bitstreams/2cbffdb9-f726-5771-8e94-50f902624f56/content

14. Chen S, Kuhn M, Prettner K, Bloom DE. The global macroeconomic burden of road injuries: estimates and projections for 166 countries. The Lancet Planetary Health. 2019 3: e390–e398. 10.1016/S2542-5196(19)30170-6.

15. Robertson LS. Reversal of the road death trend in the U.S. in 2015–2016: An examination of the climate and economic hypotheses. Journal of Transport & Health. 2018 Jun 1 [cited 2022 Aug 11];9:161–8. 10.1016/j.jth.2018.04.005

16. He L, Liu C, Shan X, Zhang L, Zheng L, Yu Y, et al. Impact of high temperature on road injury mortality in a changing climate, 1990–2019:A global analysis. Science of The Total Environment [Internet]. 2023 Jan 20;857:159369. 10.1016/j.scitotenv.2022.159369

17. Clark DE. Effect of population density on mortality after motor vehicle collisions. Accident Analysis & Prevention. 2003 Nov;35(6):965–71.

18. Yang K, Law TH, Megat-Usamah Megat-Johari, Hu Q. Economic Growth, Income Disparity, and Road Fatalities: Analyzing the Elderly-to-Younger-Adult Ratio Through Global Panel Data. Transportation Research Record Journal of the Transportation Research Board. 2025 May 29;2679(8):706–20. 10.1177/03611981251337465

19. World Population Review. Estimated Road Traffic Death Rate 2021. https://worldpopulationreview.com/country-rankings/road-deaths-by-country

20. World Health Organization. Death on the roads based on the WHO Status Report on Road Safety, 2018.

21. Global Data Lab. Geospatial Data (v0.2), 2025. https://globaldatalab.org/geos/table/

22. Populationpyramid.net. Population density. 2025. https://www.populationpyramid.net/population-density/2020/#google_vignette

23. NUMBEO.com. 2026. Cost of Living Index by Country. https://www.numbeo.com/cost-of-living/rankings_by_country.jsp?title=2021

24. National Safety Council. Injury Facts. 2026. https://injuryfacts.nsc.org/motor-vehicle/overview/age-group-comparisons/

25. William Haddon Jr, Valien P, McCarroll JR, Umberger CJ. A controlled investigation of the characteristics of adult pedestrians fatally injured by motor vehicles in Manhattan. Journal of Chronic Diseases. 1961. 14:655–678.

26. Ross HL, Campbell DT, Glass GV. Determining the social effects of a legal reform: the British “Breathalyser” Crackdown of 1967. American Behavioral Scientist. 1970. 13:493–509, https://journals.sagepub.com/doi/abs/10.1177/000276427001300402

27. H. Laurence Ross. Social Control Through Deterrence: Drinking-and-Driving Laws. Annual Review of Sociology. 1984. 10:21–35. 10.1146/annurev.so.10.080184.000321

28. Eduard Zaloshnja, Miller TR, Galbraith MS, Lawrence BA, DeBruyn LM, Bill N, Hicks, KR, Keiffer M, Perkins R. Reducing injuries among Native Americans: five cost-outcome analyses. Accident Analysis & Prevention. 2003. 35:631–639.

29. D Short, Robertson LS. Motor vehicle death reductions from guardrail installation. Journal of Transportation Engineering. 1998. 124:501–502. 10.1061/(ASCE)0733-947X(1998)124:5(501)

30. Elvik R, Høye A, Vaa T, Sørensen M, editors. The Handbook of Road Safety Measures. 2009.

31. de Andrade L, Vissoci JR, Rodrigues CG, Finato K, Carvalho E, Pietrobon R, de Souza EM, Nihei OK, Lynch C, de Barros Carvalho MD. Brazilian Road traffic fatalities: a spatial and environmental analysis. PLoS One. 2014 Jan 30;9(1):e87244. 10.1371/journal.pone.0087244.

